# Off-Trial: Real-World Weight Loss on Tirzepatide and Semaglutide

**DOI:** 10.64898/2026.07.14.26357502

**Authors:** Brian Erly, Shanmugesh Raja

## Abstract

**Background:** GLP-1 receptor agonist trials are tightly controlled: standardized titration, intensive dietary counseling, frequent in-person follow-up, and rigorous exclusion criteria. The real world is none of those things. In a U.S. telehealth GLP-1 program, diet engagement, exercise, medication choice, dose timing, and out-of-pocket cost vary substantially from patient to patient. Whether trial-level efficacy translates into the outcomes a patient and clinician will actually see is an open question, and the answer matters, because telehealth is now where most GLP-1 prescribing happens.

**Methods:** We conducted a retrospective cohort study of 13,507 adults who used a single GLP-1 agent (tirzepatide or semaglutide) through the Mochi Health telehealth obesity program and had a documented six-month weight observation. The primary outcome was achievement of ≥10% total body weight loss at six months. To address selection bias in the tirzepatide–semaglutide comparison, we used 1:1 nearest-neighbor propensity-score matching on age, sex, baseline BMI, baseline weight, and comorbid diabetes, hypertension, dyslipidemia, and prior bariatric surgery (recorded at intake), with a 0.25 SD caliper on the propensity logit. We drew a directed acyclic graph (DAG) with a clinical co-author to make the identifying assumptions explicit and to mark where unobserved variables (insurance, socioeconomic status, concomitant medications such as metformin) limit causal interpretation. We report multivariable predictors via logistic regression, compute an E-value for the matched contrast, and benchmark our point estimates against landmark RCT outcomes.

**Results:** Overall, 59.1% of patients achieved ≥10% loss at six months, with mean loss of 11.5% (median 11.3%). Threshold attainment was 86.6% at ≥5%, 59.1% at ≥10%, 27.5% at ≥15%, and 9.1% at ≥20%. The unadjusted tirzepatide–semaglutide response gap was +16.0 percentage points (68.8% vs 52.8%); after 1:1 propensity-score matching (3,480 pairs, all post-match |SMD| *<* 0.05) the gap was +18.1 percentage points (69.6% vs 51.6%, 95% CI +15.9 to +20.3). Matching on the measured covariates did not attenuate the advantage, indicating that selection on those characteristics does not explain it; the matched risk ratio was 1.35 (E-value 2.04). The gap was unchanged when a self-reported insurance indicator was added to the matching (+18.4 pp) and remained large (+14.2 pp) within patients who reached a therapeutic dose. Multivariable predictors of response were tirzepatide (OR 2.10, 1.95–2.26), female sex (OR 1.37, 1.20–1.56), and prior bariatric surgery (OR 1.36, 1.18–1.57); response was lower with comorbid diabetes (OR 0.84, 0.77–0.92) and, modestly, with higher baseline BMI per unit (OR 0.98, 0.97–0.99). Response varied by baseline BMI, from 58.0% in overweight patients (BMI *<*30) and a peak of 63.5% in Obese I to 52.4% in Obese III.

**Conclusions:** Real-world response to GLP-1 therapy in a telehealth setting is meaningfully attenuated from RCT benchmarks but remains clinically substantial: roughly three in five patients reach the 10% threshold. The tirzepatide advantage over semaglutide is large and, notably, does not shrink under propensity-score matching on measured confounders, so it is not an artifact of the observed selection variables; an unmeasured confounder would need a risk-ratio association of about 2.0 with both drug choice and response to explain it away (E-value 2.04). The findings are observational, conditional on the DAG’s identifying assumptions, and unmeasured confounders (insurance, socioeconomic status, concomitant medications) remain possible.

## 1. Introduction

GLP-1 receptor agonist trials are built to show a drug at its best. Patients enroll at academic centers, titrate on a standardized schedule, receive intensive dietary counseling and frequent in-person follow-up, are screened to exclude many real-world comorbidities, and are kept adherent by study staff. Under those conditions, semaglutide 2.4 mg/wk produces mean weight loss of approximately 14.9% over 68 weeks^1^ and tirzepatide 15 mg/wk produces 20.9% over 72 weeks.^2^

Almost nothing about ordinary care looks like that. In a U.S. telehealth GLP-1 program, the same patient might sit on a sub-therapeutic dose for protocol or tolerability reasons, might or might not engage a dietitian, might or might not log any exercise, might face a $25 copay or a $1,200 cash bill, and might be quietly self-titrating up or down based on side effects without telling the platform. Diet, exercise, medication choice, dose timing, and out-of-pocket cost all vary patient to patient. The clinically relevant question is not what a drug can do under ideal supervision but what a clinician and patient should actually expect when those constraints relax.

This paper answers that question for 13,507 adults in a national U.S. telehealth GLP-1 program, and it does so along three lines. First, we describe the full distribution of six-month weight-loss outcomes once trial-grade control is removed. Second, because patients in this setting are not randomized between tirzepatide and semaglutide, we ask whether a credible effectiveness difference between the two drugs survives adjustment for selection on observed characteristics. Third, we identify the baseline patient-level factors that independently predict reaching a clinically meaningful weight-loss threshold.

We are explicit about causal interpretation throughout. We use propensity-score matching to address selection on observed covariates, and we draw a DAG so that a reviewer can read the identifying assumptions directly. Where the DAG exposes unobserved confounders, most notably concomitant medications such as metformin, which serves both as a diabetes treatment and as a weight-loss adjunct, we report the limitation transparently and quantify robustness to it with an E-value rather than claiming to have removed it. We do not claim the matched estimate is causal in a strict bioequivalence sense. We claim it is the best observational estimate available under explicit assumptions.

## 2. Methods

### 2.1 Cohort

This is a retrospective cohort study using electronic health record and pharmacy data from Mochi Health, a U.S. telehealth obesity program operating across all 50 states. We began with 71,302 adults who had subscribed, completed intake, and used a single GLP-1 agent (tirzepatide only or semaglutide only) with an approved, dispensed first refill; patients who used both agents sequentially were excluded so that the drug contrast is not confounded by switching. Of these, 13,543 had a measured weight in the six-month follow-up window. After excluding records with implausible weights or weight changes, the analytic cohort was 13,507 patients, of whom 5,297 used tirzepatide and 8,210 used semaglutide.

### 2.2 Exposure, weights, and outcome

Drug class was assigned from the medication on the patient’s refills. Baseline weight was the measured weight closest to the first refill within a *±*30-day window, taken as the maximum across intake, refill, encounter, and patient-reported sources; where no measured baseline was available we used the intake (eligibility) weight. The six-month weight was the measured weight closest to 180 days after the first refill within a 135–225 day window. Percent total body weight loss was computed as (baseline *−* six-month) / baseline. The primary outcome was ≥10% loss at six months, consistent with FDA-accepted obesity pharmacotherapy endpoints.^3^ Secondary outcomes were mean percent loss and achievement of the ≥5%, ≥15%, and ≥20% thresholds.

### 2.3 Causal framework and DAG

The causal target is the difference in six-month ≥10% response between tirzepatide and semaglutide had each patient counterfactually received each drug. To make the identifying assumptions explicit we drew the DAG in Figure 1 with a clinical co-author using dagitty.^5^ Adjusted variables are observed and entered the propensity model or the outcome regression. Insurance is only coarsely observed (a self-reported intake item) and is added in a sensitivity analysis (Section 3.6); socioeconomic status and concomitant medications such as metformin are unobserved. Their direct effects on the exposure are partially absorbed by observed children such as BMI, weight, diabetes, age, and sex, but they cannot be fully blocked.

**Figure 1:**
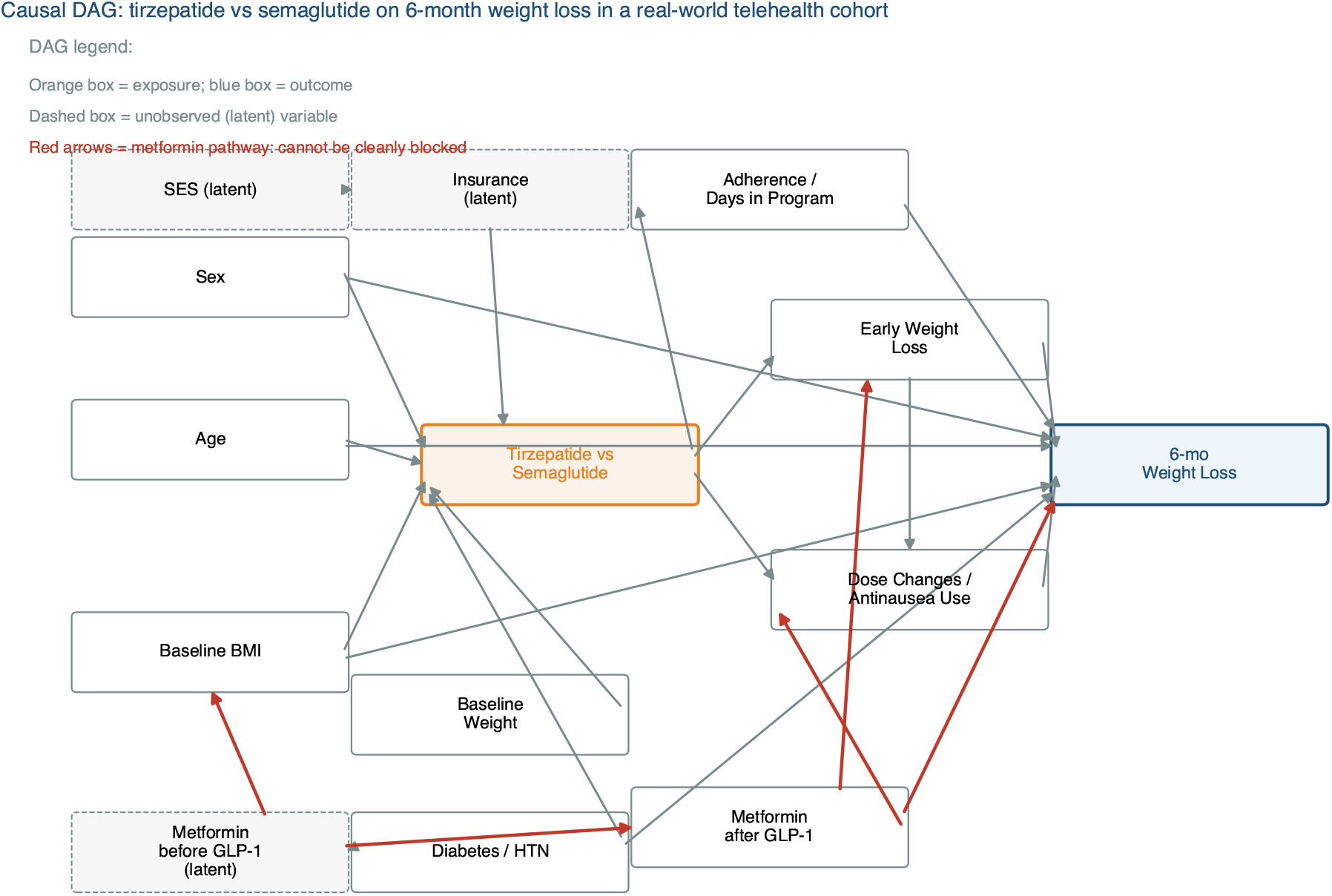
Causal DAG: tirzepatide vs semaglutide on six-month weight loss in a real-world telehealth cohort. The orange box is the exposure; the blue box is the outcome. Dashed boxes are unobserved (latent) variables. The metformin pathway (shown in red) is an example of an unmeasured medication confounder: metformin is taken by some patients for diabetes (a confounder) and by others as a weight-loss adjunct (a mediator), so it cannot be cleanly adjusted away. Metformin use was not reliably ascertainable in our intake data; we treat it, together with insurance and socioeconomic status, as a source of residual confounding and quantify robustness with an E-value (Section 4.3).

### 2.4 Propensity-score matching

We performed 1:1 nearest-neighbor matching without replacement, with a 0.25 SD caliper on the logit propensity score.^4^ The propensity model was a logistic regression of drug class (tirzepatide = 1, semaglutide = 0) on age, sex, baseline BMI, baseline weight, and comorbid diabetes, hypertension, dyslipidemia, and prior bariatric surgery, the last four recorded as intake diagnoses. Balance was assessed by standardized mean differences (SMD), with |SMD| *<* 0.10 considered adequate. Matching yielded 3,480 tirzepatide–semaglutide pairs; the matched estimand is the average effect in the matched (overlap) population. We computed an E-value for the matched risk ratio to quantify how strong an unmeasured confounder would have to be to explain the result.^8^

### 2.5 Multivariable analysis

We fit a logistic regression of the binary ≥10% loss outcome on drug class and the propensity-model covariates, reporting odds ratios with 95% confidence intervals.

### 2.6 Sensitivity analyses

We ran two sensitivity analyses for the tirzepatide–semaglutide contrast. First, because insurance is plausibly correlated with both drug choice and response, we repeated the matching with a self-reported insurance-coverage indicator added to the propensity model. Second, because achieved dose and adherence sit on the causal path between molecule and outcome (the drug works partly by being tolerated and titrated to a therapeutic level), we did not adjust for them in the primary model, but we report them descriptively by arm (refill count as an adherence proxy; whether the patient reached a therapeutic dose, defined as tirzepatide ≥5 mg or semaglutide ≥1.0 mg) and stratify the response gap by therapeutic-dose attainment. The dose-stratified contrast conditions on a mediator and therefore estimates a different quantity than the primary total-effect comparison; we report it only to show how much of the advantage operates through reaching a therapeutic dose.

### 2.7 What we deliberately did not analyze, and why

We did not split the comparison further by branded versus compounded formulation. That question has a separate confounding structure (insurance, SES) and is addressed in a companion cost study. Mixing the tirzepatide-versus-semaglutide and branded-versus-compounded questions in a single comparison produces estimates that mean neither thing clearly, so we keep them separate. We also did not analyze adverse-event rates; our outcome is weight loss, and safety requires different exposure denominators and adverse-event ascertainment than our data support. Finally, we did not model concomitant medications (e.g., metformin) or antiemetic use, because neither could be reliably ascertained from the available intake data; we treat them as unmeasured confounders (Section 4.3) rather than report unreliable estimates.

### 2.8 Software

Analyses were performed in Python 3.12 with scikit-learn 1.9, pandas 2.3, statsmodels 0.14, and SciPy 1.17. The matched-difference confidence interval was obtained by 2,000-replicate bootstrap.

## 3. Results

### 3.1 Cohort and overall outcomes

Among 13,507 patients, mean six-month weight loss was 11.5% (median 11.3%). Threshold attainment rates are shown in Figure 2. A clear majority reached the ≥10% threshold (59.1%); the ≥15% threshold was reached by 27.5%, and ≥20% by 9.1%. These rates fall meaningfully below those seen in landmark RCTs, where SURMOUNT-1 reported ≥15% in approximately 56% of patients on tirzepatide 15 mg.^2^ The gap is the expected attenuation when titration is variable, follow-up is asynchronous, and behavioral engagement is heterogeneous. Baseline characteristics by drug are shown in Table 1.

**Table 1:** Table 1. Baseline characteristics by drug. Values are mean (SD) unless noted.

| Characteristic | Tirzepatide (n = 5,297) | Semaglutide (n = 8,210) |
| --- | --- | --- |
| Age, years | 44.7 (11.5) | 41.8 (11.7) |
| Female, % | 87 | 89 |
| Baseline BMI, kg/m<sup>2</sup> | 37.3 (7.0) | 36.5 (6.7) |
| Baseline weight, lb | 229 (50) | 222 (47) |
| Mean six-month loss, % | 13.0 (6.5) | 10.6 (6.1) |

**Figure 2:**
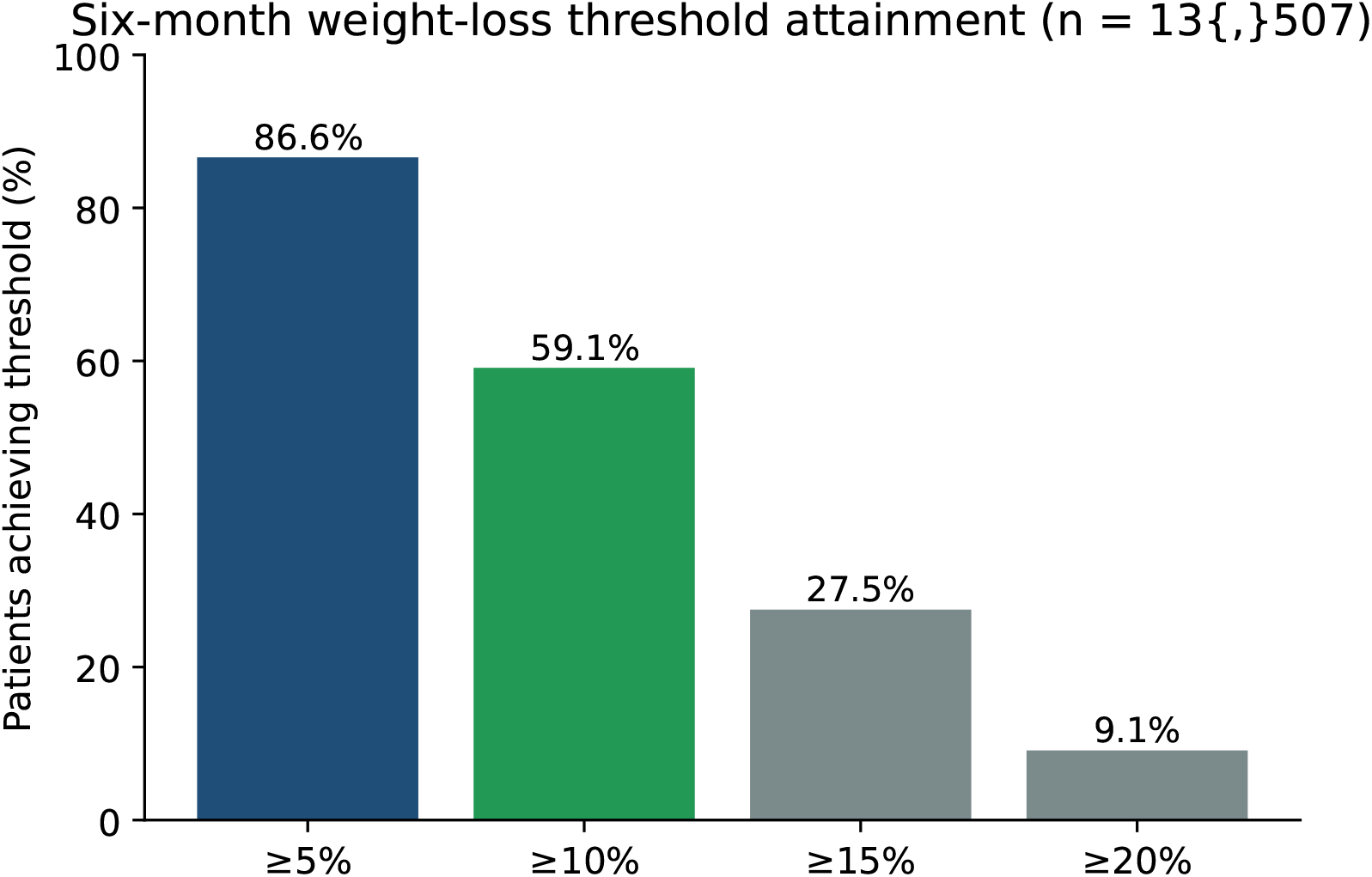
Distribution of six-month weight-loss outcomes by threshold. Bars show the percentage of the 13,507-patient cohort reaching each weight-loss threshold at six months. Rates are below landmark RCT rates but remain clinically substantial: a majority of patients reach ≥10% loss.

### 3.2 Tirzepatide vs semaglutide: unadjusted vs PSM-matched

In the unadjusted comparison, tirzepatide showed a 16.0 percentage point higher ≥10% response rate (68.8% vs 52.8%; Figure 3A). After 1:1 propensity-score matching on age, sex, BMI, weight, comorbid diabetes, hypertension, dyslipidemia, and prior bariatric surgery, the gap was 18.1 percentage points (69.6% vs 51.6%, 95% CI +15.9 to +20.3, 3,480 pairs). Matching achieved good balance on every covariate (Table 2); the largest pre-match imbalance, in age (SMD 0.25), was reduced to *−*0.03 after matching, and all matched SMDs were below 0.05.

**Table 2:** Table 2. Covariate balance before and after matching (standardized mean differences).

| Covariate | SMD before matching | SMD after matching |
| --- | --- | --- |
| Age | 0.247 | -0.031 |
| Female sex | -0.067 | 0.015 |
| Baseline BMI | 0.119 | 0.025 |
| Baseline weight | 0.143 | 0.025 |
| Comorbid diabetes | 0.068 | 0.034 |
| Hypertension | 0.039 | -0.040 |
| Dyslipidemia | 0.075 | 0.015 |
| Prior bariatric surgery | 0.037 | 0.018 |

**Figure 3:**
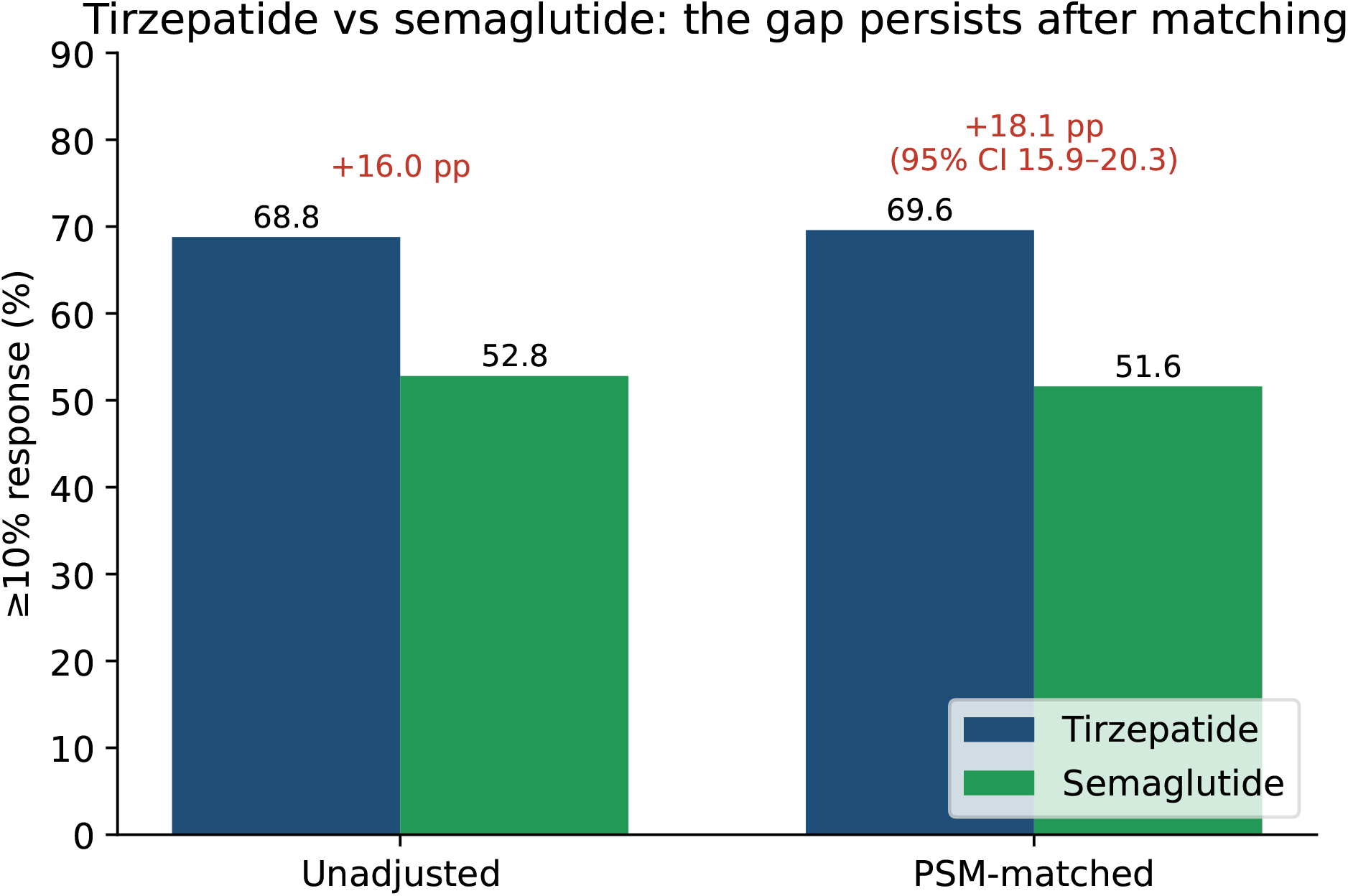
Tirzepatide vs semaglutide effectiveness. Six-month ≥10% response rate in the unad-justed cohort (left pair of bars) and in the 1:1 PSM-matched sample (right pair). The advantage does not shrink with matching (16.0 pp unadjusted; 18.1 pp matched, 95% CI 15.9–20.3), indicating that selection on the measured covariates does not account for it.

The central finding is that matching did *not* attenuate the tirzepatide advantage: the gap was 16.0 pp unadjusted and 18.1 pp in the matched sample. Selection on the measured characteristics therefore does not explain the difference. This makes the advantage more robust to measured confounding than a naive read of an observational comparison would suggest; the matched risk ratio was 1.35, with an E-value of 2.04, meaning an unmeasured confounder would have to be associated with both drug choice and ≥10% response by a risk ratio of about 2.0 to explain the result away.

### 3.3 Multivariable predictors

Table 3 reports the multivariable odds ratios for ≥10% loss. After adjustment, tirzepatide carried an OR of 2.10 (1.95–2.26). Female sex was associated with higher response (OR 1.37), as was prior bariatric surgery (OR 1.36). Comorbid diabetes was associated with lower response (OR 0.84), and higher baseline BMI carried a small per-unit reduction in the odds of reaching the relative ≥10% threshold (OR 0.98 per unit); hypertension and dyslipidemia were not significantly associated with response.

**Table 3:**
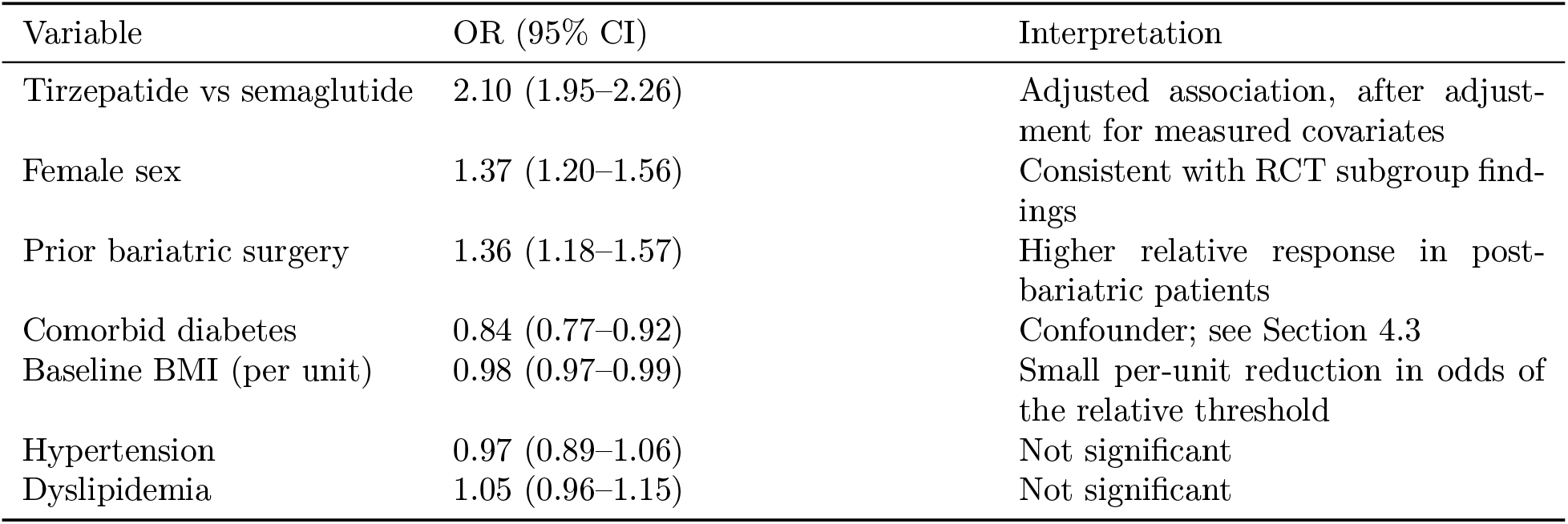
Table 3. Multivariable odds ratios for ≥10% loss at six months.

The diabetes coefficient should not be read as a treatment effect. In the DAG, diabetes is a confounder rather than a mediator; the OR captures the difference between diabetic and non-diabetic patients, not the effect of diabetes status on a fixed patient. The negative per-unit BMI coefficient reflects the relative (percent) outcome: higher-BMI patients lose more absolute weight but a slightly smaller *percent* of a larger starting weight, and the relationship is non-monotone across BMI categories (Section 3.4).

### 3.4 Subgroup heterogeneity by baseline BMI

Response varied across baseline BMI categories (Figure 4). Patients with BMI *<*30 (overweight) reached ≥10% loss at 58.0%, rising to a peak of 63.5% for Obese I (30–35), then declining to 59.6% for Obese II (35–40) and 52.4% for Obese III (40+). The decline in the highest-BMI group likely reflects both ceiling effects on percent loss off a larger denominator and behavioral heterogeneity in that group.

**Figure 4:**
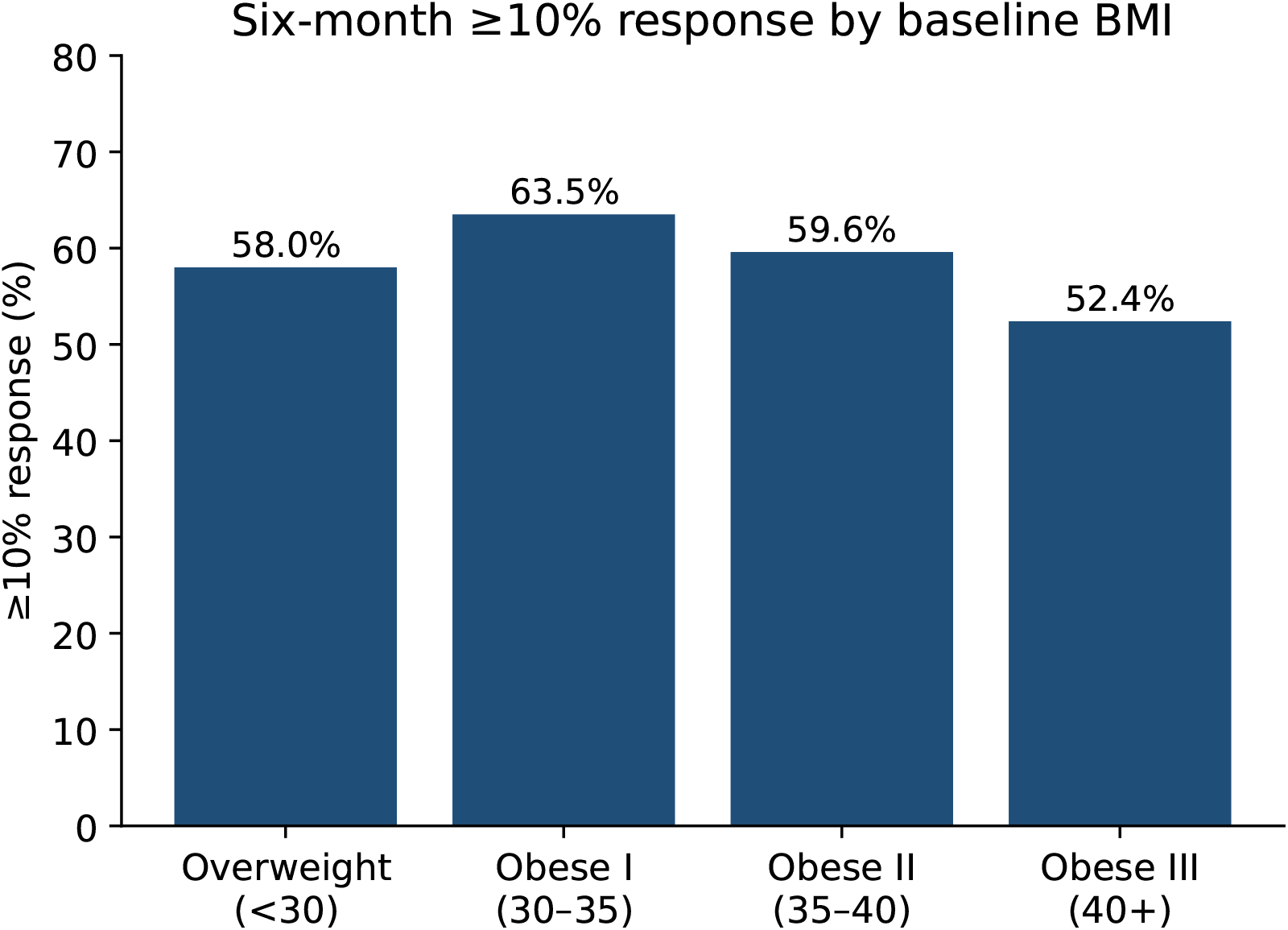
Response rate by baseline BMI category. Percentage of patients reaching ≥10% loss at six months stratified by baseline BMI category. Response peaks in Obese I and declines in Obese III.

### 3.5 Sensitivity analyses

The tirzepatide advantage was robust to the two sensitivity analyses (Table 4). Self-reported insurance coverage was modestly higher among tirzepatide patients (7.9% vs 5.1%), consistent with the selection concern, but adding the insurance indicator to the propensity model left the matched gap essentially unchanged at +18.4 pp (95% CI +16.1 to +20.7). On the mediator pathway, tirzepatide patients refilled slightly more often (mean 9.2 vs 8.3 refills) and were far more likely to reach a therapeutic dose (98.5% vs 87.6%). Yet conditioning on therapeutic-dose attainment did not collapse the gap: among patients who reached a therapeutic dose the advantage was +14.2 pp (69.0% vs 54.8%), and among those who did not it was +15.0 pp (53.8% vs 38.8%). Reaching a therapeutic dose more often therefore accounts for only a small part of the tirzepatide advantage; most of it persists within dose-equalized strata.

**Table 4:** Table 4. Sensitivity analyses for the tirzepatide–semaglutide ≥ 10% response gap. Dose-stratified rows condition on a mediator and estimate a different quantity than the primary total-effect contrast.

| Analysis | Tirzepatide | Semaglutide | $\Delta$ (pp) [95% CI] |
| --- | --- | --- | --- |
| Primary PSM (8 covariates) | 69.6% | 51.6% | +18.1 [15.9, 20.3] |
| PSM + self-reported insurance | — | — | +18.4 [16.1, 20.7] |
| Reached therapeutic dose (n = 12,411) | 69.0% | 54.8% | +14.2 |
| Below therapeutic dose (n = 1,096) | 53.8% | 38.8% | +15.0 |

## 4. Discussion

### 4.1 What this paper says, in plain language

In a U.S. telehealth GLP-1 cohort where diet, exercise, dose timing, and out-of-pocket cost all vary patient to patient, roughly three in five patients reach the 10% weight-loss threshold at six months, a magnitude of loss that in randomized trials of semaglutide has translated into reduced cardiovascular events in patients with overweight or obesity but without diabetes.^6^ The mean loss of 11.5% sits meaningfully below the RCT benchmarks, but it is the right number to ground patient expectations: an unselected patient walking into the clinic should expect about that, with substantial variation around it. Tirzepatide was associated with a substantially higher ≥10% response rate than semaglutide, and, unusually for an observational contrast, that advantage did not shrink under matching on the measured confounders.

### 4.2 Why response is below RCT benchmarks

The RCT comparison should not be read as “telehealth underperforms.” The trial setting is engineered to maximize the drug’s effect through standardized titration to the maximum tolerated dose, intensive in-person dietary counseling, and screening out of the comorbidities most likely to attenuate response; STEP-3, which paired semaglutide with 30 visits of intensive behavioral therapy, reached ≥15% loss in 55.8% of participants, illustrating how much the behavioral scaffold contributes in the trial setting.^7^ The real-world setting has none of that by design. Many patients in our cohort are not on the maximum dose at six months because they could not tolerate it, did not titrate up, or could not afford to. The comparison is also timepoint-mismatched: our outcomes are measured at six months, whereas the STEP-1 (68 weeks) and SURMOUNT-1 (72 weeks) benchmarks are read out at roughly 16–17 months, by which point weight loss is still accruing, so part of the apparent attenuation reflects the shorter observation window rather than a true effectiveness shortfall. Calibrating expectations to a partial-dose, partial-engagement reality is the relevant clinical task, and 11.5% mean loss is the right anchor for it.

### 4.3 What we cannot claim, and why

The matched tirzepatide–semaglutide gap of 18.1 pp is our best observational estimate, not a bioequivalence statement. That it does not shrink under matching tells us the measured covariates do not explain it, but it cannot rule out unmeasured confounding, and three sources remain.

#### Concomitant medications are unobserved

Metformin in particular is taken by some patients for diabetes (a confounder) and by others as a weight-loss adjunct (a mediator), so it cannot be cleanly adjusted even when measured. We could not reliably ascertain it from intake data and did not model it; it is therefore part of the residual confounding the E-value bounds rather than a variable we controlled.

#### Insurance is coarsely observed; SES is not

Tirzepatide patients reported insurance coverage slightly more often (7.9% vs 5.1%), but adding that indicator to the matching did not move the gap (Table 4), so the observed insurance signal does not explain the advantage. True socioeconomic status (income, education) is not collected, and the self-reported coverage item is coarse, so a finer insurance/SES pathway could still contribute.

#### Adherence and achieved dose are mediators, not confounders

We deliberately did not adjust for refill counts or achieved dose, because both sit on the causal path from molecule to outcome; the matched estimate is a total-effect contrast between molecules as used in practice, not a dose-equalized comparison. The dose-stratified sensitivity shows the advantage persists within dose-equalized strata (+14.2 pp among patients reaching a therapeutic dose), so tirzepatide’s edge is only partly explained by its higher rate of reaching maintenance dosing. The E-value of 2.04 quantifies the joint strength an unmeasured confounder would need to overturn the result.

### 4.4 The branded-versus-compounded question is not addressed here

We deliberately did not split this comparison further by branded versus compounded formulation. That comparison has its own confounding structure (insurance access, socioeconomic status, adherence), and treating it as a subgroup nested inside the tirzepatide-versus-semaglutide analysis muddles both estimates. We address branded versus compounded separately in a companion study that uses a different DAG and a different propensity model.

### 4.5 Limitations

#### Single-platform retrospective

The cohort is one telehealth provider’s patients. External validation in independent telehealth cohorts and in traditional obesity-medicine settings is needed before generalizing the magnitude of the tirzepatide–semaglutide advantage.

#### Selection on six-month observation

Patients with a documented six-month weight may be more engaged than the full prescribed cohort, and we did not formally address differential ascertainment between drug arms; the analytic cohort (13,507) is the subset of single-agent users with a measured follow-up weight.

#### Observational, not bioequivalence

See Section 4.3.

#### Unmeasured confounders

Insurance, SES, and concomitant medications are not directly observed; the E-value quantifies robustness but does not remove the possibility.

#### Subgroup analyses are descriptive

Subgroup analyses are reported without formal interaction testing, and we did not correct for multiple comparisons.

## 5. Conclusions

In 13,507 adults who used tirzepatide or semaglutide through a U.S. telehealth GLP-1 program, six-month weight loss was meaningfully attenuated from RCT benchmarks but remained clinically substantial: 59.1% reached ≥10% loss, with mean loss of 11.5%. Tirzepatide was associated with a markedly higher ≥10% response rate than semaglutide (unadjusted +16.0 pp; matched +18.1 pp, 95% CI +15.9 to +20.3), and the advantage did not shrink under propensity-score matching on measured confounders, so it is not an artifact of the observed selection variables (E-value 2.04). The result is observational, conditional on the DAG’s identifying assumptions, and does not address the branded-versus-compounded question, which is addressed in a companion study. For real-world expectation-setting at intake, the 11.5% mean and 59.1% threshold-attainment figures are the right anchors, not the RCT means.

## Data Availability

Individual patient-level data are private and cannot be shared publicly. De-identified data and the analysis code can be provided upon reasonable request to the corresponding author.

## Author Contributions

**Brian Erly:** Conceptualization, DAG development, Supervision, Writing, Clinical validation. **Shanmugesh Raja:** Conceptualization, Data curation, Formal analysis, Methodology, Software, Visualization, Writing.

## Ethics Approval

This study is a secondary analysis of de-identified data collected during routine clinical care at Mochi Health. As the analysis involves no direct patient contact, no intervention, and uses only data de-identified per HIPAA Safe Harbor [45 CFR 164.514(b)(2)], it was determined to not constitute human subjects research as defined by 45 CFR 46.102. Formal IRB review was therefore not required.

## Conflicts of Interest

S.R. and B.E. are contractors for Mochi Health, the telehealth provider that supplied the de-identified data analyzed in this study. Mochi Health had no role in the study design, the analysis, the interpretation of results, or the decision to submit the work for publication; the authors retained full independent control over all of these. No external funding was received.

## Notes

### Author Declarations

This study is a secondary analysis of de-identified data collected during routine clinical care at Mochi Health. As the analysis involves no direct patient contact, no intervention, and uses only data deidentified per HIPAA Safe Harbor [45 CFR 164.514(b)(2)], it was determined to not constitute human subjects research as defined by 45 CFR 46.102. Formal IRB review was therefore not required.

